# Comprehensive assessment of Age-Specific Mortality Rate and its incremental changes using a composite measure: A sub-national analysis of rural Indian women

**DOI:** 10.1101/2022.04.25.22274281

**Authors:** Divya Sharma, Tanvi Kiran, KP Junaid, Vineeth Rajagopal, Saraswati Sharma

## Abstract

**Background:** Diverse socio-economic and cultural issues contribute to adverse health outcomes and increased mortality rates among rural Indian women across different age categories. The present study aims to comprehensively assess age-specific mortality rates and their temporal trends using a composite measure at the sub-national level for rural Indian females to capture cross-state differences.

**Methods:** A total of 19 states were included in the study to construct a composite age-specific mortality index for 2011 (base year) and 2018 (reference year) and examine the incremental changes in the index values across these years at the sub-national level in India. Sub-index values were calculated for each component age group and were subsequently used to compute the composite ASMR index using the geometric mean method. Based on the incremental changes, the performance of states was categorized into four different typologies.

**Results:** Improvement in mortality index scores in the 0-4 years age group was documented for all states. The mortality rates for the 60+ age group were recorded to be high for all states. Kerela emerged as the overall top performer in terms of mortality index scores, while Bihar and Jharkhand were at the bottom of the mortality index table. The overall mortality composite score has shown minor improvement from base year to reference year at all India level.

**Conclusions:** An overall reduction in the mortality rates of rural Indian women has been observed over the years in India. The success of public health interventions to reduce the under-five mortality rate is evident as the female rural mortality rates have reduced sizably for all states. Nevertheless, there is still sizable scope for reducing mortality rates for other component age groups. Additionally, there is a need to divert attention toward the female geriatric (60+ years) population as the mortality rates are still high.

## Introduction

Nearly 65% of the Indians predominantly reside in rural areas [1], and half of them fall below the poverty line [2]. Several studies conducted in developing countries have indicated the presence of a strong association between low socio-economic status, poor health, and inaccessible health care facilities [3]. Due to the unfavorable socio-economic conditions, the rural population faces a recurring struggle for survival and succumbs to it most of the time [4]. Disparities in the life span of rural and urban inhabitants have also been documented by many studies globally [5–7]. It can be explained by the disproportionate distribution of public resources toward healthcare in urban and rural areas, thus increasing the gap in terms of healthcare facilities [8]. The low investment in the health infrastructure of rural areas leads to management issues, a shortage of dedicated cadre of the health workforce, and insufficient training of healthcare workers, which contribute to poor health services at large. The other factors accounting for poorer survival of rural residents include inefficient illness management, remote and inaccessible healthcare facilities, lack of preventive and screening measures, and low levels of knowledge and awareness [7].

Significant health inequalities and disparities in health status are also seen within India, leading to differences in the mortality rates of vulnerable populations like women and children. These inequalities affect populations at the national and sub-national levels [9]. The discrepancies in the health status resulting in high mortality are majorly reported in the Empowered Action Group (EAG) states of India, which are socioeconomically backward and lag in the demographic transition [10]. Bihar, Madhya Pradesh, Jharkhand, Chhattisgarh, and Uttar Pradesh are the few EAG states and are usually found at the bottom of the tables in most of the developmental parameters [11]. The major reasons for the poor performances of these states, specifically in terms of health outcomes and high mortality rates, include poverty, overpopulation, low literacy levels, gender discrimination, and poor health infrastructure [12].

Though biological differences exist between the genders however [13]; the health outcome differences and inequalities are more evident in the case of rural Indian women. Women face diverse socio-economic and cultural issues attributed in terms of gender discrimination, such as female infanticide, child marriage, dowry, domestic violence, lack of education, and unavailability of proper sanitation and healthcare facilities [14–16]. These indicators contribute to adverse health outcomes and increased mortality rates among rural women across different age categories [10]. Various targeted public health programmes have been implemented to reduce mortality rates, especially in the EAG states. Age-specific interventions have also been launched to improve nutrition among women [17] and reduce mortality, particularly among infants and under-five children [18]. The programmes have successfully reduced mortality to a great extent, but the progress has been uneven. There is still a striking difference between the age-specific mortality rates at the sub-national level in India, especially concerning women residing in rural areas [19,20].

There is a gap in the literature on assessing differences in the Age-Specific Mortality Rates (ASMR) pertaining to rural women across different Indian states. To the best of our knowledge, no published study in India has been conducted to quantify and transform the age-specific mortality values into a single composite value at the sub-national level, especially for rural females. The present study addresses the lacuna by constructing age-specific mortality index for four major component age groups and a composite ‘Age-specific mortality index’ for rural Indian women at the sub-national and national levels for base and reference years to examine the temporal changes, if any. The proposed composite measure includes mortality values of all component age groups within its ambit to capture India’s cross-state differences. In this backdrop, the specific objectives of the study can be defined as follows; a) to comprehensively quantify the mortality values at the sub-national level in India by constructing a composite age-specific mortality index for rural Indian women using a rigorous methodology involving historical benchmark values for mortality; b) to examine the incremental changes in mortality index scores from base year to reference year at the sub-national level.; c) to map and categorize Indian states into not improved, least, moderately and highly improved classification based on the incremental changes of the mortality index scores. Thus, examining the state-specific and age-specific determinants leading to sizable mortality rates among rural Indian women is essential to address the health inequalities. The identification of determinants necessitates the assessment of age-specific mortality rates and their temporal trends in quantifiable terms at the sub-national level for rural Indian females, which forms the very rationale and aim of the present study.

## Methods

### A. Data source

The secondary data on age-specific mortality rates of states was extracted from Sample Registration System (SRS) statistical reports available on the web portal of the office of the Registrar General and Census Commissioner under the Indian Ministry of Home Affairs. The Census of India generates data on population statistics, including vital statistics and census [21]. A total of 19 states have been included in the study to construct a composite age-specific mortality index for 2011 (base year) and 2018 (reference year) and examine the incremental changes in the index values across these years at the sub-national level in India.

### B. Components of Age-Specific Mortality Rate

Age-Specific Mortality Rate (ASMR) is a mortality rate confined to a particular age group [22]. The SRS statistical reports publish ASMR by sex and residence for four-component age groups, i.e., 0-4 years, 5-14years, 15-59 years, and 60 years and above. Since the present study aimed to construct a composite ASMR index for rural females, the data pertaining to all the aforementioned component age groups of rural females were extracted for the years commencing from 1971 to 2018 for the identification of benchmark values required for data normalization.

### C. Data cleaning

The extracted data was randomly chosen and rechecked to assess errors in data entry. Followed by that, the data was checked for the missing values. Out of 28 Indian states, the data was completely missing for 7 states, bringing the tally down to 21 states. Subsequently, data for two Indian states, namely; Uttarakhand, was available after 2014 and for Telangana for the year 2018 only. The historical values of these states were not available in the literature and thus, these states were excluded from the study.

### D. Computation of composite Age-specific Mortality Index

#### i. Data normalization

The extracted ASMR data was normalized based on the standardized procedure by employing the ‘Minimum-Maximum approach’ used by international and national agencies and organizations to compute various composite indices [4-5]. Since the indicators in the study were negative in nature, where an actual lower value means better performance, therefore the values were normalized and scaled using the following formula [23]: -

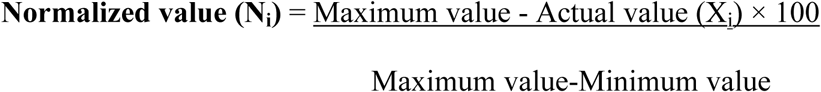

The minimum and maximum values refer to the benchmark values identified using the following procedure.

#### ii. Identification of ‘minimum and maximum’ (benchmark values) using historical data

The Census of India has been publishing annual SRS statistical reports since 2011, before which these reports were published intermittently commencing from 1971. The benchmark values were identified using historical ASMR values published in SRS statistical reports from 1971 to 2018 for each component age group. The historical data, being a crucial tool in statistical procedures, is the data often disaggregated or in different measuring [25] units that have been preserved for a long period [26] as it reflects information having substantial significance. Historical data is mainly required to identify the thresholds, which are then used for data normalization. For negative indicators like mortality, the maximum thresholds should not be obtained from the data range for which the index has to be calculated (2011 to 2018 in the present study) using the geometric mean approach [27,28]. The rationale governing this principle is that despite having non-zero (positive) values for the component/domains of the composite index, the entire index is rendered to be zero in the event of a single domain registering a zero value (which is transformed through normalization process explained above). The minimum and maximum values (benchmarks) are given in the supplementary table **(S1 Table)**.

#### iii. Construction of composite Age-specific Mortality Index

The normalized value computed for the four component age groups reflects the sub-index values for these age groups, which were subsequently used to compute the composite ASMR index using the geometric mean method. The geometric mean method has the upper hand over other measures as it is less altered by extreme values, is robust and thus maintains an overall balance in data distribution. Further, it is a standardized technique for index construction adopted by reputed international agencies/organizations, including United Nations Development Programme (UNDP) [24,29]

The index scores range from 0-to 100 for each indicator. A higher index score (closer to 100) indicates a lower ASMR value, and a lower index score (closer to zero) indicates a higher ASMR value.

### E. Incremental changes and classification of states into different typologies

Incremental change measures the change in the index score from 2011 (base year) to 2018 (reference year). The highest positive incremental change across all the component age groups was 13.9, reported in the 60+ category. Based on the incremental changes, the states are divided into four groups: ‘Not improved’ (<=0 incremental change), ‘Least improved’ (0.01 to 3 points increase), ‘Moderately improved’ (3.01 to 6 points increase), and ‘Highly improved’ (> 6 points increase). The categorization was done using the percentile method (P_20_ and P_40_ determining cut off points for least improved and moderately improved, respectively) and is based on the approach followed by the National Institution for Transforming India (NITI Aayog-the apex public policy ‘think tank’ of the Government of India) for categorizing health index scores.

### F. Data sharing and Ethical Considerations

The study is purely based on secondary data, which is freely available to the general public. The study does not involve any interaction with the participants/human subjects. Hence, ethical approval was not required for this study. Further, the excel files depicting the normalized values and computed composite index score for age-specific mortality rates for rural Indian women across different Indian states have already been uploaded to the Mendeley data repository [30] (https://data.mendeley.com/datasets/v5gskzj78x/1) and can be easily accessed by the interested users/researchers/ academicians and alike.

## Results

### Descriptive characteristics of component age groups of ASMR for rural Indian women

The means of AMSR values of the four component age groups for each state were calculated for a span of 8 years (2011-2018) **(Table 1)**. At the all-India level, the highest mean death rate (41.63±0.42) for rural Indian females was observed for the 60+ age group, while the lowest was for the 5-14 years age group (0.15±0.08). In three out of four component age groups, barring 60+ years, Kerela recorded the lowest mean death rate values for rural Indian women.

**Table 1:** Descriptive characteristics of component age groups of Age-specific mortality rates of rural Indian women at the sub-national and national levels from 2011-2018.

|  | Mean± SD |  |  |  | Coefficient of variation (%) |  |  |  |
| --- | --- | --- | --- | --- | --- | --- | --- | --- |
| States | Death rates (rural females) |  |  |  | Death rates (rural females) |  |  |  |
|  | 0-4 years | 5-14 years | 15-59 years | 60+ years | 0-4 years | 5-14 years | 15-59 years | 60+ years |
| Andhra Pradesh | 10.51±0.87 | 0.49±0.22 | 3.34±0.18 | 39.04±1.43 | 8.25 | 44.45 | 5.53 | 3.66 |
| Assam | 16.18±2.17 | 0.90±0.38 | 3.30±0.49 | 46.76±4.51 | 13.43 | 42.00 | 14.93 | 9.63 |
| Bihar | 11.66±1.51 | 0.74±0.24 | 2.35±0.3 | 53.48±5.55 | 12.99 | 32.36 | 15.26 | 10.37 |
| Chhattisgarh | 13.10±1.65 | 0.83±0.39 | 3.76±0.47 | 49.33±5.94 | 12.56 | 47.50 | 12.46 | 12.04 |
| Gujarat | 11.63±1.64 | 0.65±0.15 | 2.51±0.17 | 32.70±2.66 | 14.08 | 23.26 | 6.87 | 8.12 |
| Haryana | 11.61±1.11 | 0.56±0.38 | 2.45±0.19 | 36.65±2.94 | 9.56 | 67.15 | 7.87 | 8.03 |
| Himachal Pradesh | 7.86±1.96 | 0.45±0.25 | 1.88±0.35 | 33.48±3.05 | 24.88 | 55.71 | 18.86 | 9.12 |
| Jammu&<br>Kashmir** | 8.51±1.85 | 0.55±0.18 | 2.14±0.21 | 27.64±5.19 | 21.77 | 32.23 | 9.98 | 18.76 |
| Jharkhand | 11.33±1.86 | 0.98±0.63 | 3.05±0.42 | 50.88±5.08 | 16.45 | 64.34 | 13.69 | 9.98 |
| Karnataka | 8.49±0.94 | 0.45±0.18 | 3.11±0.32 | 44.44±2.18 | 11.08 | 39.40 | 10.37 | 4.91 |
| Kerala | 2.79±0.47 | 0.15±0.08 | 1.56±0.34 | 34.16±3.28 | 16.77 | 50.40 | 21.89 | 9.61 |
| Madhya Pradesh | 17.70±2.94 | 1.14±0.36 | 2.88±0.32 | 46.16±2.23 | 16.60 | 31.85 | 11.12 | 4.83 |
| Maharashtra | 5.75±0.41 | 0.38±0.10 | 2.66±0.50 | 38.04±3.65 | 7.14 | 27.60 | 18.72 | 9.59 |
| Odisha | 14.28±2.08 | 1.00±0.38 | 3.19±0.30 | 41.94±3.43 | 14.58 | 37.80 | 9.55 | 8.17 |
| Punjab | 7.78±1.94 | 0.35±0.19 | 2.73±0.34 | 36.34±2.99 | 24.90 | 52.90 | 12.37 | 8.23 |
| Rajasthan | 14.91±2.67 | 0.70±0.24 | 2.16±0.23 | 34.89±1.35 | 17.91 | 34.15 | 10.47 | 3.88 |
| Tamil Nadu | 5.13±0.61 | 0.48±0.16 | 2.93±0.26 | 39.10±2.13 | 11.97 | 33.29 | 8.91 | 5.45 |
| Uttar Pradesh | 16.76±2.46 | 1.01±0.20 | 3.46±0.41 | 45.05±3.85 | 14.68 | 19.35 | 11.85 | 8.54 |
| West Bengal | 7.05±1.05 | 0.50±0.21 | 2.31±0.19 | 41.59±2.51 | 14.95 | 42.76 | 8.15 | 6.04 |
| **India** | **12.11±1.60** | **0.75±0.12** | **2.83±0.12** | **41.63±0.42** | **13.21** | **15.94** | **4.12** | **1.02** |

\*\* J&K was an Indian state till 2019 and is now administered as a union territory by India

In the age groups 0-4 years and 5-14 years, Madhya Pradesh reported the highest mean value (17.70±2.94 and 1.14±0.36), while Kerela (2.79±0.47 and 0.1 5±0.08) registered the lowest mean death rates for the rural Indian females. Chhattisgarh recorded the highest (3.76±0.47) and Kerala the lowest mean death rates (1.56±0.34) in 15-59 years of age. In the 60+ age group, Bihar documented the highest mean value (53.48±5.55), while Jammu & Kashmir (J&K) reported the lowest mean death rate (27.64±5.1). Furthermore, estimates of the coefficient of variation showed the highest variation in the 5-14 years age group and lowest variation in the 60+ years age group.

### Age-specific Mortality Index scores of component age groups and its comparison between the base year and reference year for rural Indian women

The study computed sub-indices for mortality with regard to each component age group and compared their index scores for the base year (2011) and reference year (2018) **(Table 2 & 3)**. Compared to the base year, index scores of mortality of rural women for the age group of 0-4 years increased for all the Indian states. Kerala accounted for the highest index score for 0-4 years in the base year (98.8) and in the reference year (99.8), and Madhya Pradesh documented the lowest index scores (80.5 and 87.5) in both the years. In terms of the incremental change between the base year and reference year for the age group 0-4 years, the highly improved (HI) states with the highest incremental changes include Madhya Pradesh (7.0) and Rajasthan (7.0). In contrast, the least improved states with the lowest incremental changes were Maharashtra (0.5), Kerala (1.0), and Tamil Nadu (1.6) (**Fig. 1)**.

**Table 2:** Incremental changes in mortality Index scores of age groups 0-4 and 5-14 years for rural Indian women from the base year to reference year.

|  | 0-4 years |  |  | 5-14 years |  |  |
| --- | --- | --- | --- | --- | --- | --- |
| State | Index score Base year (2011) | Index score Reference Year (2018) | Incremental change | Index score Base year (2011) | Index score Reference Year (2018) | Incremental change |
| Andhra Pradesh | 90.9 (9) | 92.7 (10) | 1.8 (LI) | 86.2 (7) * | 93.8 (3) * | 7.6 (HI) |
| Assam | 83.5 (17) | 89.1 (16) | 5.6 (MI) | 89.2 (5) * | 96.9 (1) | 7.7 (HI) |
| Bihar | 88.9 (10) | 93.1 (8) | 4.2 (MI) | 81.5 (9) * | 87.6 (7) | 6.1 (HI) |
| Chhattisgarh | 87.2 (14) | 91.3 (13) | 4.1 (MI) | 80.0 (10) | 93.8 (3) * | 13.8 (HI) |
| Gujarat | 87.9 (12) | 92.8 (9) | 4.9 (MI) | 87.7 (6) * | 92.3 (4) * | 4.6 (MI) |
| Haryana | 88.8 (11) | 92.1 (12) | 3.3 (MI) | 93.8 (3) | 95.3 (2) | 1.5 (LI) |
| Himachal Pradesh | 92.5 (5) | 97.0 (3) | 4.5 (MI) | 96.9 (1) | 93.8 (3) * | -3.1 (NI) |
| Jammu & Kashmir** | 91.0 (8) | 95.8 (6) | 4.8 (MI) | 95.4 (2) * | 92.3 (4) * | -3.1 (NI) |
| Jharkhand | 87.4 (13) | 92.3 (11) | 4.9 (MI) | 87.7 (6) * | 92.3 (4) * | 4.6 (MI) |
| Karnataka | 92.2 (6) | 94.4 (7) | 2.2 (LI) | 92.3 (4) | 90.7 (5) * | -1.6 (NI) |
| Kerala | 98.8 (1) | 99.8 (1) | 1.0 (LI) | 96.9 (1) | 96.9 (1) | 0 (NI) |
| Madhya Pradesh | 80.5 (19) | 87.5 (18) | 7.0 (HI) | 86.2 (7) * | 90.7 (5) * | 4.5 (MI) |
| Maharashtra | 96.0 (2) | 96.5 (4) * | 0.5 (LI) | 95.4 (2) * | 93.8 (3) * | -1.6 (NI) |
| Odisha | 85.1 (15) | 90.7 (15) | 5.6 (MI) | 81.5 (9) * | 92.3 (4) * | 10.8 (HI) |
| Punjab | 91.5 (7) | 96.5 (4) * | 5.0 (MI) | 89.2 (5) * | 92.3 (4) * | 3.1 (MI) |
| Rajasthan | 83.8 (16) | 90.8 (14) | 7.0 (HI) | 84.6 (8) | 89.2 (6) | 4.6 (MI) |
| Tamil Nadu | 95.9 (3) | 97.5 (2) | 1.6 (LI) | 89.2 (5) * | 95.3 (2) * | 6.1 (HI) |
| Uttar Pradesh | 81.7 (18) | 87.7 (17) | 6.0 (MI) | 81.5 (9) * | 86.1 (8) | 4.6 (MI) |
| West Bengal | 93.3 (4) | 96.3 (5) | 3.0 (LI) | 87.7 (6) * | 95.3 (2) * | 7.6 (HI) |
| **India** | **87.6** | **92.1** | **4.5 (MI)** | **86.2** | **90.7** | **4.5 (MI)** |

'NI' = Not Improved ( $\leq 0$ ); 'LI' = Least Improved (0-3); 'MI' = Moderately Improved (3.01-6); 'HI' = Highly Improved (>6.01)
The ranks for each Indian state have been displayed in the parentheses of the table. \* Denotes tied ranks.
\*\* J&K was an Indian state till 2019 and is now administered as a union territory by India.

**Fig. 1:**
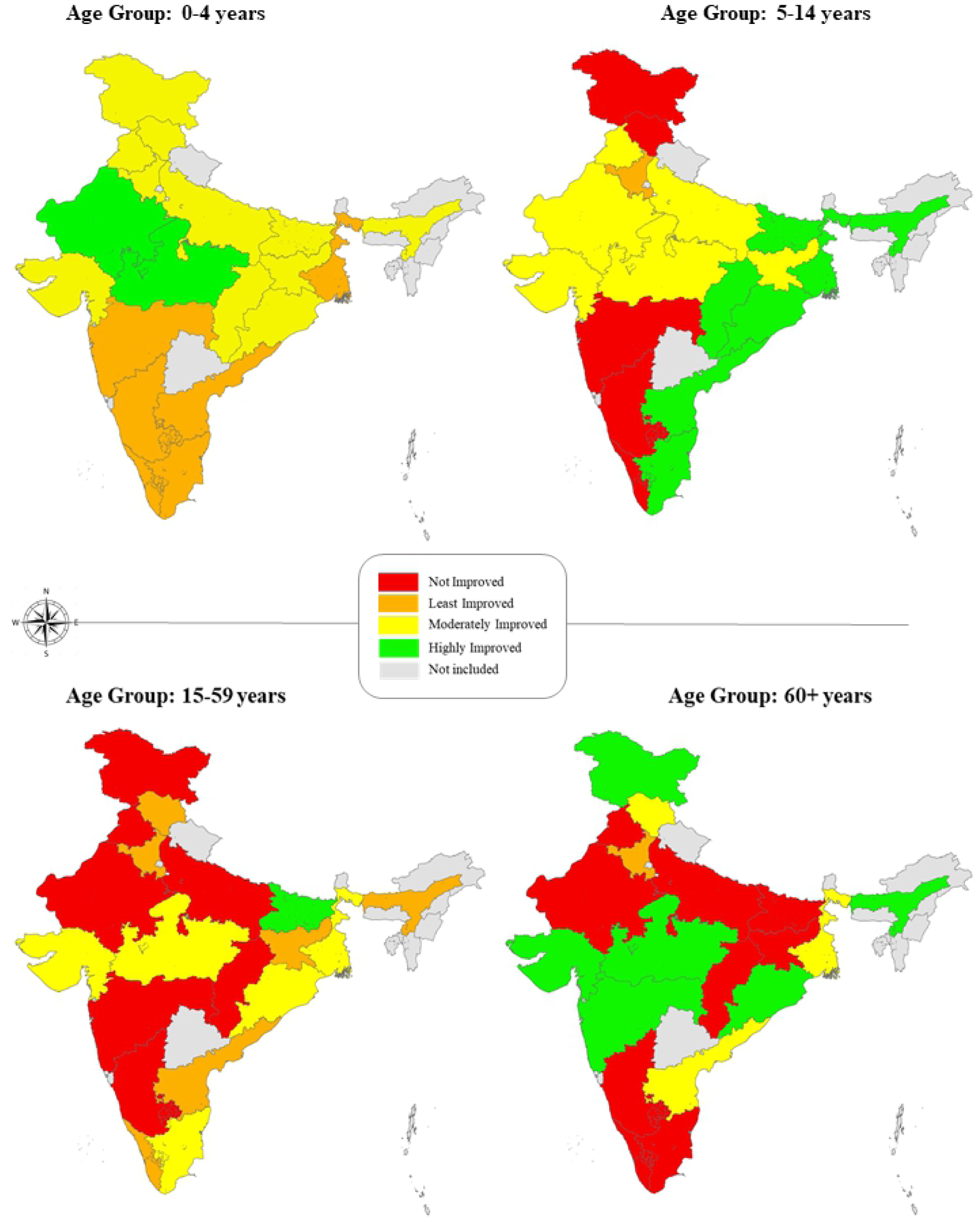
Mapping of Indian states based on incremental changes in index scores for each component age group.

In the age group 5-14 years, the index scores of states were relatively higher than other component age groups. However, there was a decline in the mortality index score for states that registered high index values in the base year. Fourteen states reported an increase in the mortality score in the reference year for this particular age group. A remarkable improvement was seen in Chhattisgarh (13.8) and Odisha (10.8) (**Fig. 1)**. Meanwhile, Himachal Pradesh (−3.1), Jammu & Kashmir (−3.1), Karnataka (−1.6), and Maharashtra (−1.6) recorded negative changes in the mortality scores.

For the 15-59 age group **(Table 3)**, the mortality score of rural Indian women ranged from 68.0 to 95.1. Kerela reported the highest index scores in base (93.4) and reference years (95.1), while Chhattisgarh reported the lowest scores in both years (76.2 & 68.0). There was no change in Karnataka, Maharashtra, and Rajasthan index scores from base year to reference year. Based on the incremental changes, Bihar showed the highest improvement (8.2), while Odisha (5.0) and Madhya Pradesh (4.9) improved moderately in the age category 15-59 years (**Fig. 1)**. The states with negative changes in the mortality scores included Chhattisgarh (−8.2), Punjab (−4.9), Jammu & Kashmir (−1.6), and Uttar Pradesh (−1.6)

**Table 3:**
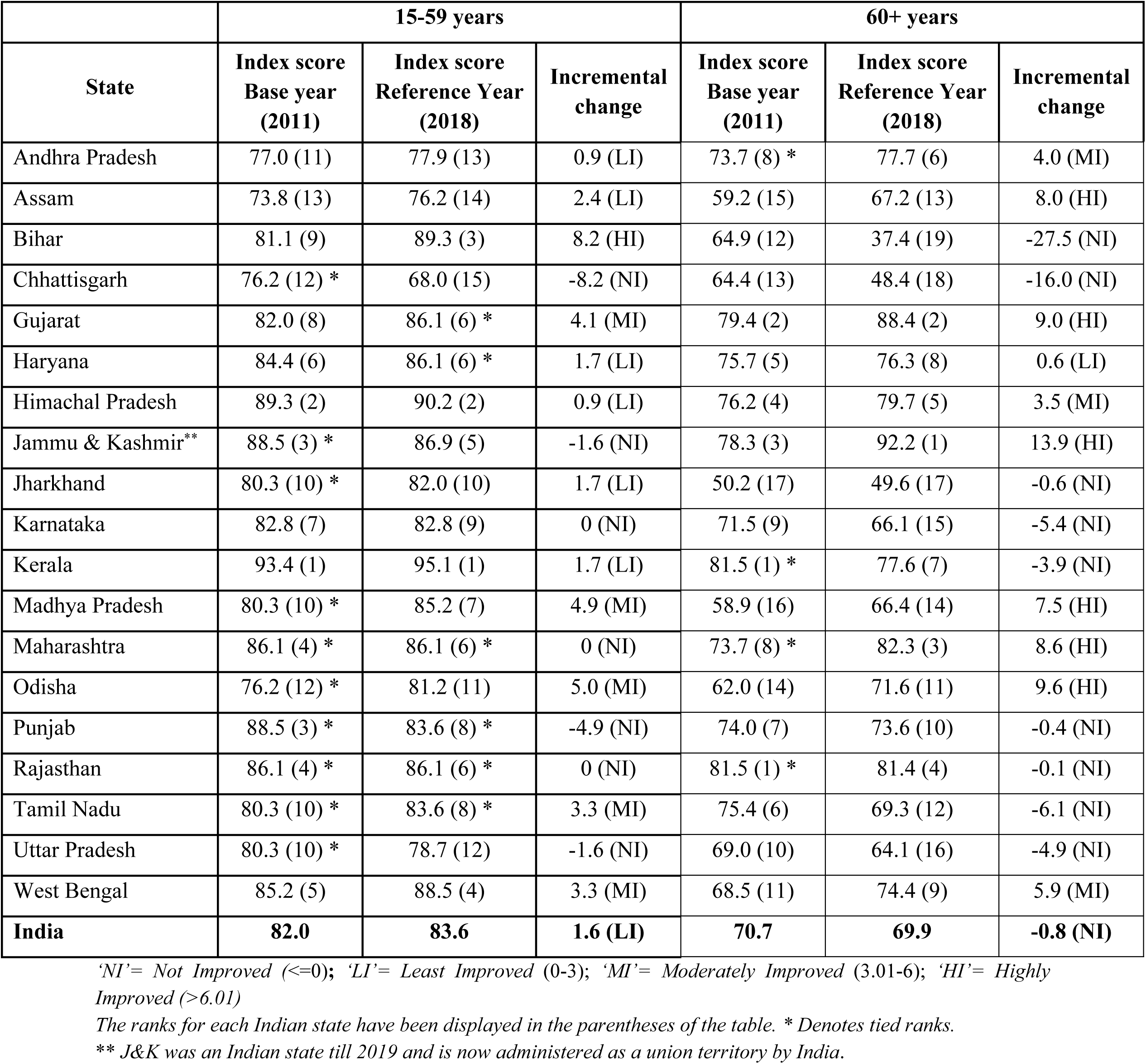
Incremental changes in mortality Index scores of age groups 15-59 and 60+ years for rural Indian women from the base year to reference year.

A wide disparity of mortality scores for rural women was seen in the 60+ years age group ranging from 37.4 to 92.2 in the reference year. Kerela (81.5) registered the highest index score in the base year and Jammu & Kashmir (92.2) in the reference year. Jharkhand (50.2) and Bihar (37.4) recorded the lowest index scores in the base year and reference year in this particular age group. Ten states showed improvement in index scores from base year to reference year. Jammu & Kashmir recorded the highest improvement (13.9) in this age group. It was followed by Odisha (9.6) and Gujarat (9.0) (**Fig. 1)**. In contrast, Bihar (−27.5), Chhattisgarh (−16.0), and Tamil Nadu (−6.1) documented notable negative changes in the mortality index values for this age group. At all India level, the mortality scores sizably increased for both 0-4 (4.5) and 5-14 years age groups (4.5), whereas it slightly improved and decreased for 15-59 (1.6) and 60+ (−0.8) age groups, respectively for rural women.

### Composite Age-specific Mortality Index (ASMI) scores of component age groups and its comparison between the base year and reference year for rural Indian women

A composite age-specific mortality index for rural Indian women was developed by aggregating sub-indices for all component age groups using the geometric mean method **(Table 4)**. The highest mortality index score in the base year was 92.4, while the lowest was 74.6. In the reference year, the index score range was 72.3 to 91.9. Kerala reported the highest composite index scores in the base year (92.4) and reference (91.9). Jharkhand (74.6) and Bihar (72.3) documented the lowest composite index scores in base and reference years, respectively. Fifteen states recorded an increase in the index scores, out of which three states improved highly, four reported moderate improvement, and eight made least improvements in the mortality scores **(Fig. 2)**.

**Table 4:**
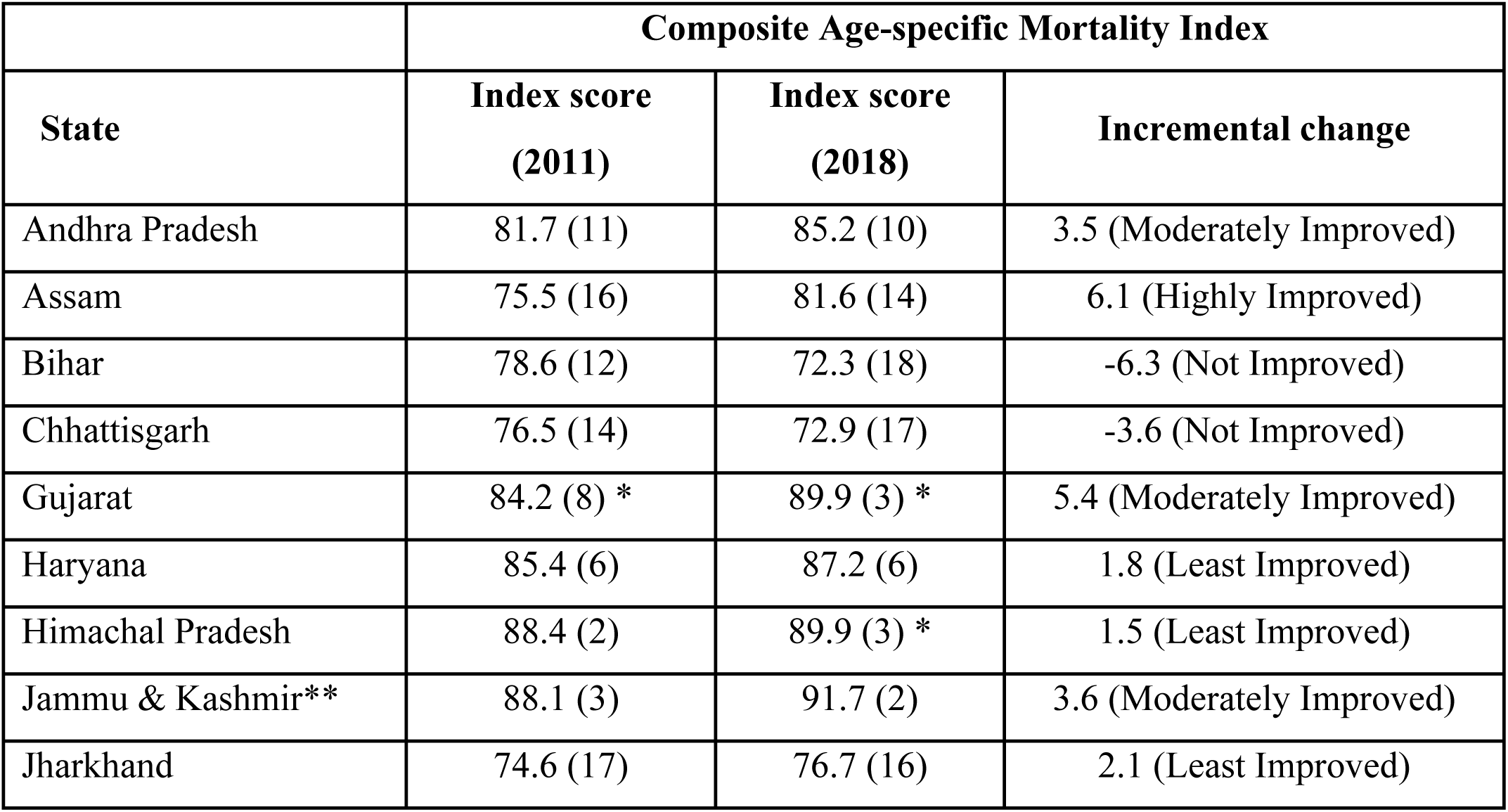
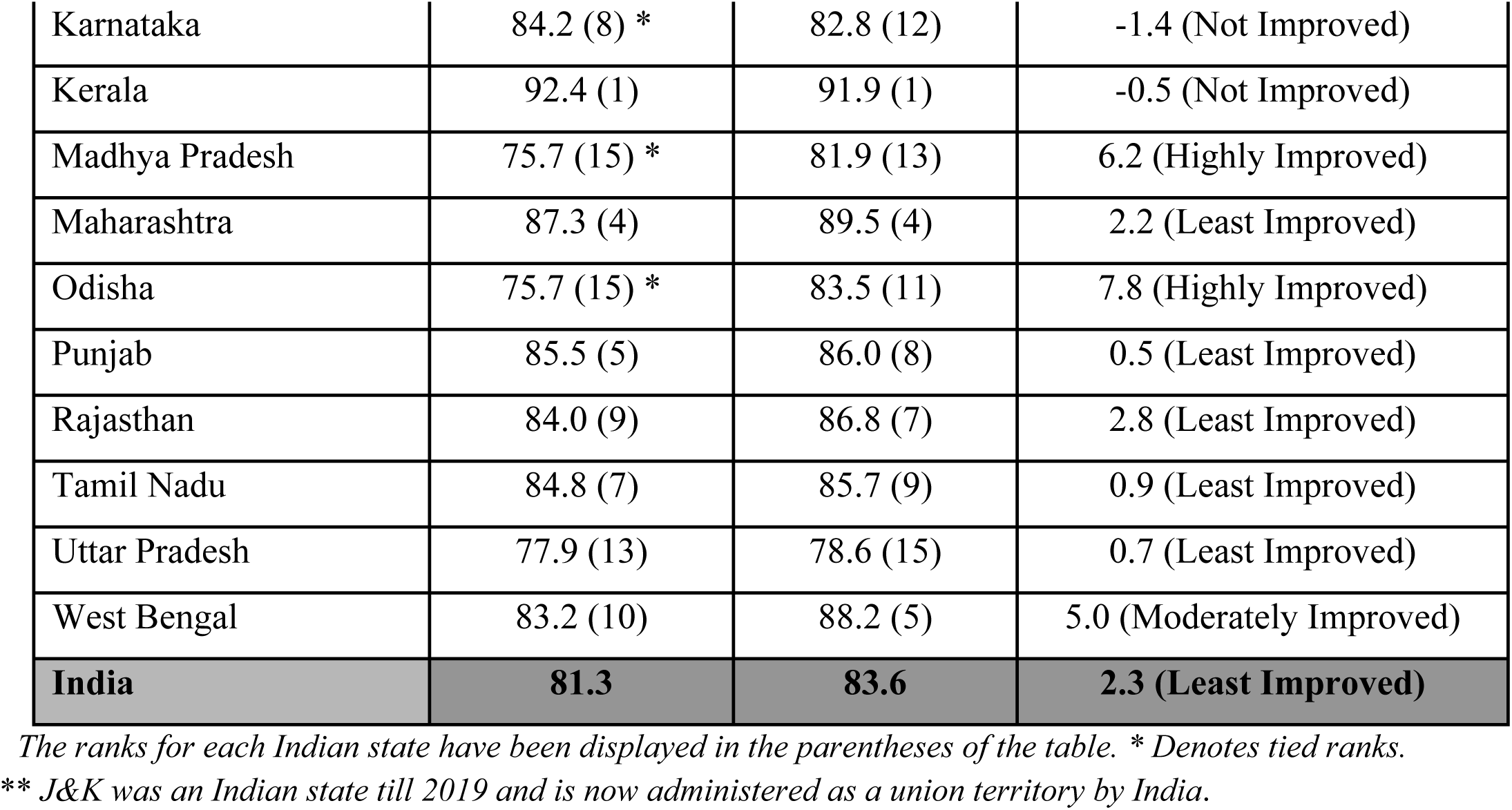
Composite Age-specific Mortality Index scores for rural Indian women and its Incremental changes from the base year to reference year.

**Fig. 2:**
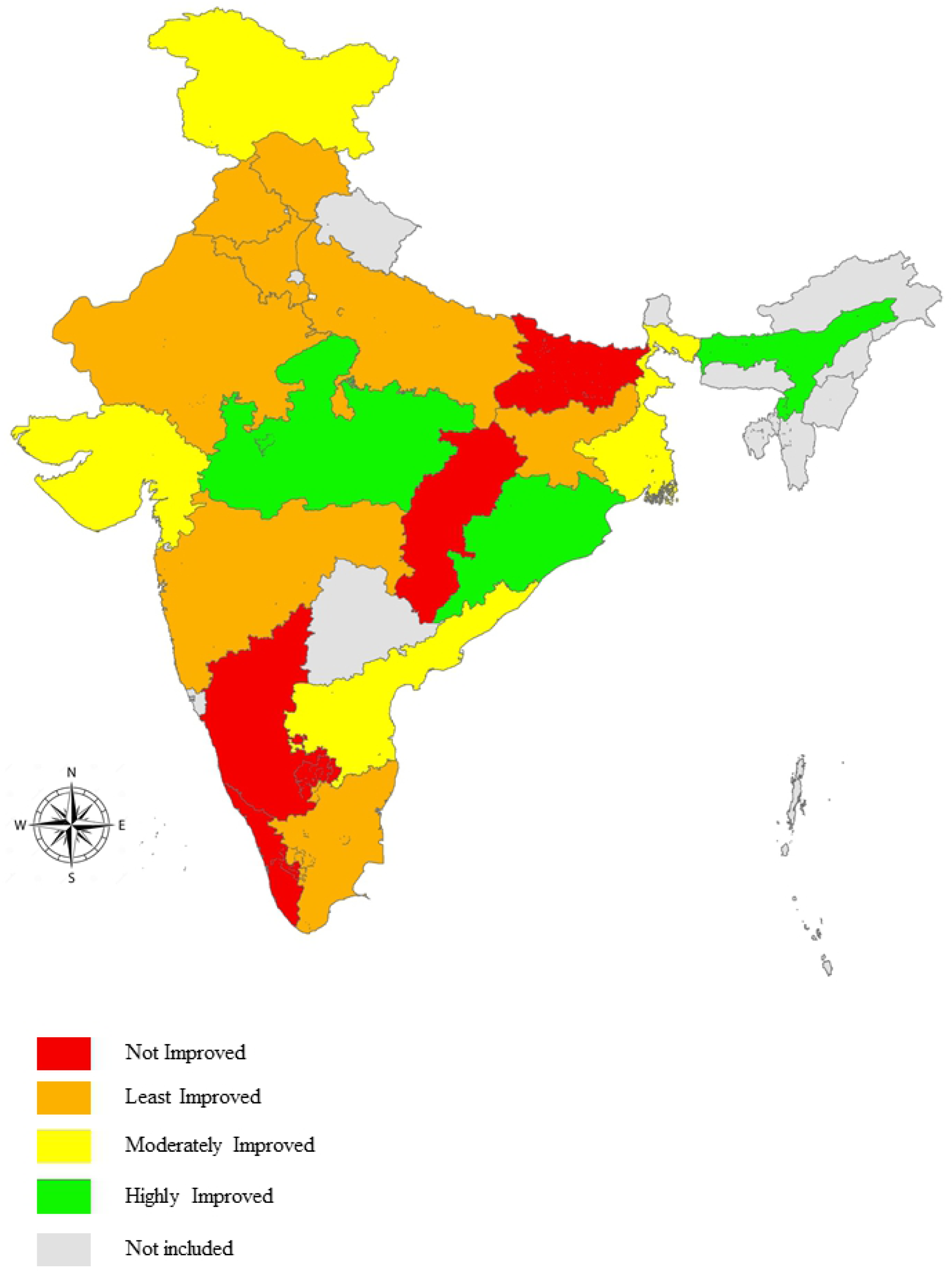
Mapping of Indian states into typologies based on incremental changes of composite Age-Specific Mortality Index scores.

### Top and bottom performing Indian states based on Incremental changes in composite Age-Specific Mortality Index scores for rural Indian women

Odisha reported the highest incremental change (7.8) from base year to reference year. Substantial improvements were also observed in Madhya Pradesh (6.2) and Assam (6.1). On the contrary, composite ASMI scores of Bihar (−6.3), Chhattisgarh (−3.6), Karnataka (−1.4), and Kerela (−0.5) reported a decline. At all India level, the incremental change in the composite mortality index score for rural women has witnessed a slight improvement (2.3). The incremental changes in the composite index are depicted in **Fig. 3**. The top five performing states **(Fig. 3)** with regard to composite mortality scores for rural Indian women have more or less remained the same, except that West Bengal has replaced the state of Punjab in the reference year **(S2 and S3 map)**. The bottom five performing states included Chhattisgarh, Assam, and Jharkhand in base and reference years. Madhya Pradesh and Odisha in the base year were replaced by Uttar Pradesh and Bihar in the reference year **(Fig. 3) (S2 and S3 map)**.

**Fig. 3:**
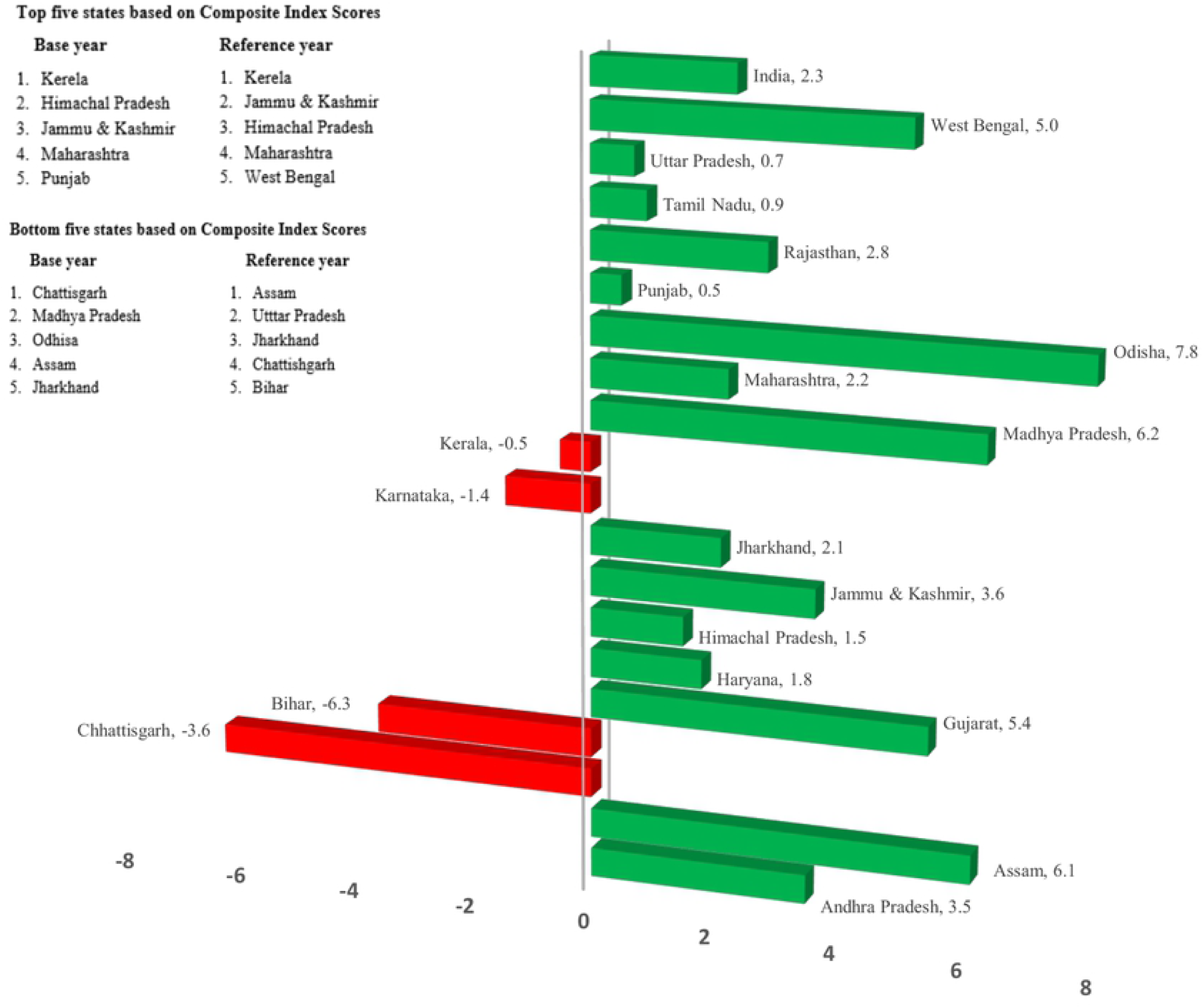
Positive and Negative Incremental changes in composite Age-Specific Mortality Index scores along with top and bottom performing states for rural Indian women.

## Discussion

The study computed mortality index scores for four major component age groups and a composite Age-specific mortality index for rural Indian women at the sub-national and national levels. Rural Indian women face several difficulties due to gender disparity and discrimination. Issues concerning lack of educational opportunities, financial inclusion, proper medical care, hygiene, and sanitation are widespread in rural areas and are encountered more by women than men [14,15]. Incremental changes in the index scores from the base to reference years have also been calculated. Based on the incremental changes, the performance of states was categorized into four levels for each component age group.

Out of the four age groups, the highest mean death rate was observed for the 60+ age group, while the lowest was for 5-14 years. The increase in mortality with age can be attributed to many factors. Irrespective of gender, the possibility of getting a chronic disease and disability increases with age, immunity, and strength also dwindle gradually, thus making the elderly more vulnerable to health problems and increasing the risk of mortality [31–33]. Further, studies on treatment-seeking behaviour in India revealed that the large majority of the elderly rural Indian population has unmet healthcare needs, thereby further substantiating the high mean death rates for elderly females as revealed by the present study [34]. Most deaths in the age group 5-14 years generally result from communicable diseases. However, a rapid decline in mortality due to infectious diseases in this age group has contributed to lower death rates [35]. Further, a study revealed that females have better survivorship in the under-five age group in India [36].

Looking at the sub-national level, the state of Madhya Pradesh documented the highest mean death rates among the states in the age groups 0-4 and 5-14 years, which can be attributed to women’s low literacy and the common practice of child marriage, especially in the rural areas of this state. The state is among the backward states and trails behind the rest of India with regard to the demographic transition [10]. Low literacy leads to a lack of awareness and affects the utilization of Antenatal Care (ANC) services [37,38]. The prevalence of child marriage being interlinked with low literacy levels is significantly associated with abortions, premature deliveries, and low birth weight infants resulting in high child mortality [38].

Chhattisgarh topped mean death rates among rural females in the 15-59 years age group. It has been reported that 42% of deaths caused by malaria in India are from Chhattisgarh alone [39]. Rural populations, particularly females, were relatively at higher risk than their male counterparts as far as malaria mortality figures are concerned in this state [40]. The maternal mortality rates are also comparatively high in the state, thus adding to death rates in the 15-59 age group [39].

In the 60+ age group, Bihar documented the highest mean death rate value. Bihar is one of India’s most populous and poorest states. It is usually placed at the lower end of all major Indian states as far as most socio-economic development parameters are concerned [11]. The prevalence and incidence of anemia were considerably high among females [41], particularly in the geriatric population [42]. With a lack of proper infrastructure and health services, the elderly population in rural areas succumb to these illnesses due to the inaccessibility of good healthcare services [43].

Jammu & Kashmir (J&K) reported the lowest mean death rate in the 60+ years. An increase in life expectancy in J&K has been noted from 1990 to 2016 [44]. In comparison to males, the life expectancy for females increased considerably by 10.86 years during the said period [44]. The determinants contributing to increased life expectancy include better healthcare delivery services, improved health infrastructure, development of supply chain mechanisms, and quality medical education in the state [45,46].

The study calculated the sub-indices for all component age groups. The index scores for 0-4 years of all states increased from base year to reference year. It indicates that the mortality rates for the age group 0-4 decreased from 2011 to 2018, which suggested better performance as far as death rates in this early age group are concerned. This trend can be attributed to the reduction in the under-5 mortality rate (U5MR), which has noticeably reduced from 66 deaths to 41 rural female deaths per 1000 live births [47] from 2011 to 2018 in India. Further, the neonatal mortality rate (NMR) has reportedly gone down by 38%, almost during a similar time period. The identifiable reason behind this is the strengthened immunization programme incorporating an umbrella of vaccines targeted to prevent fatal health outcomes[10,48,49].

Kerala was the top mortality index scorer for all component age groups in both years except for 60+ years in the reference year. Similar outcomes were seen for mean death rates, where Kerela reported the lowest mean for all age groups barring 60+ years. According to the Health Index report (2019-2020) of the National Institution for Transforming India (NITI) Aayog, which is the Indian government’s apex think tank, Kerela emerged as the top performer on multiple health-related indicators among the larger states [50]. The state’s performance in recording lower rural female death rates can be associated with various economic and health-related factors. Kerela has registered enormous improvement in income level in the last decade [51], and it presently falls under the high per capita income and growth rate category [52]. The health infrastructure of Kerela has shown extensive expansion between 2005-2019, and the state has witnessed a considerable increase in the number of Community Health Centres (CHCs) in rural areas. The number of allopathic doctors at the Primary Healthcare Centres (PHCs) has also increased significantly [12]. To add to it, it is a front runner in female literacy rates [53] and maternal education in India [54]. In context to the present study results, Kerela registered high mortality index scores, but the incremental change from base to reference year for each age group has been negligible [50]. The reason could be the low mortality rates have already reached the saturation level, and only marginal reduction is possible [55,56].

All Indian states recorded an improvement in the female mortality index scores for the 0-4 age group. Therefore, despite having a noticeable appreciation in the index score from the base to the reference year, Madhya Pradesh documented the lowest index score for both years. The health index report by NITI Aayog also reported deterioration in performance by the state for NMR and U5MR [50]. The states of Himachal Pradesh, Jammu & Kashmir, Karnataka, and Maharashtra, with high index scores in the base year, showed a decline in the reference year for the 5-14 age group. A plausible explanation for this trend is that these states had recorded relatively high mortality index scores in the reference year and had considerably reduced mortality to a great extent for this age group. Subsequently, most public health interventions and programs were directed toward reducing female mortality in other age groups [57].

In the age group 15-59, Chhattisgarh reported the lowest index score for the base year, further decreasing in the reference year. A decline in three of four health outcomes has also been witnessed for Chhattisgarh in the NITI Aayog report [50]. Similarly, for the age group 60+ years, both Chhattisgarh and Bihar witnessed a worsening of their performance in the reference year, as revealed by their respective substantial decremental changes in female mortality index scores. These states are poverty-stricken states, with most poverty concentrated in their rural areas [51]. Huge gender disparities in education levels have been recorded [58,59], wherein both these states are plagued with low female literacy levels, resulting in mass poverty [58]. The states’ health infrastructure is weak, with an acute shortage of doctors at PHCs [12]. Poverty makes the population more vulnerable to diseases, and the lack of healthcare services to address health problems results in high mortality rates.

The composite Age-specific Mortality Index (ASMI) exhibited results consistent with the sub-indices. In terms of overall index scores, the top five performers included Kerela, Himachal Pradesh, Jammu & Kashmir, Maharashtra, and Punjab in the base year. Other studies pertaining primarily to the overall health index have reported results on similar lines [23,50]. West Bengal replaced Punjab in the reference year in the top five performers. The deterioration of performance by Punjab in the age group 15-59 might have caused the change in overall rank. It could be a consequence of high maternal mortality in the region [60]. Punjab is also among the high HIV incidence states leading to higher female deaths in the state [60]. Another study reported weakened performance of Punjab in terms of a downward shift in various health outcomes [23].

The bottom five performing states were Chhattisgarh, Assam, Jharkhand, Madhya Pradesh, and Odisha in the base year. Chhattisgarh performed poorly for age groups 15-59 and 60+years and thus showed a decline in overall score from base year to reference year. In contrast, Bihar and Uttar Pradesh documented a decline in the performances in the reference year and entered the bottom five performing states category, replacing the states of Odisha and Madhya Pradesh, which recorded highly improved incremental performance in terms of composite female rural mortality scores. There was a drastic decline in the index scores of Bihar in 60+ years, causing an impact on the overall performance. The NITI Aayog health index also reported a decline in Bihar’s performance and a downward transition in all health outcomes, including institutional deliveries, Total Fertility Rates (TFR), Low Birthweight (LBW), success in treating tuberculosis, and healthcare facilities and services [23]. Uttar Pradesh had also recorded decremental changes in index scores for both 15-59 and 60+years, resulting in its entry into the bottom five performing states in the reference year.

Kerela has remained at the top of its game, thereby emerging as the best-performing state in terms of overall rural female mortality scores in both the base and reference years. The probable reasons for this consistent performance have already been discussed earlier in this study. Additionally, the overall mortality composite score has shown minor improvement from base year to reference year at all India level. It indicates that there is still room for improvement as the composite score achieved by India (83.6.) in the reference year is still noticeable points away from the maximum potential (100) achievable.

The current study faces a few limitations. Due to the non-availability of data for all states, 19 out of 28 states were included in the study, leaving the scope for future studies to be conducted incorporating the left-over states as and when the data is made available by the Indian government. Since the data on age-specific mortality groups (0-4, 5-14, 15-59, 60+) from 1971 to 2001 was disaggregated into various small age groups (5-9, 10-14, 15-19, etc.), the average values of the death rates were taken to concise them into four major component age groups to identify the threshold values for data normalization. Therefore, these threshold values for specific component age groups need to be used with caution in similar future research work.

Nevertheless, the present study has some strengths to offer. The ‘composite Age-specific Mortality Index’ is a pioneer endeavor to comprehensively quantify the mortality levels pertaining to rural Indian women at the sub-national level. The scope of usage of this unique ‘composite index’ is vast as it provides concise information from the complex and extensive data, which is more convenient to communicate and report for policy plans. The index summarizes the overall performances of the states and performances specific to component age groups, thus highlighting those age groups which need attention from the policymakers and the government. This will facilitate the reduction in mortality rates of rural Indian women at the national level as well. Further, the study has categorized the Indian states into different typologies (not improved, least/moderately/highly improved) based on their temporal performance from base to reference year. It will facilitate the laggard states to initiate customized policies/interventions to follow the performance trajectory of the frontrunner states in terms of lower rural female mortality scores. Furthermore, the threshold minimum and maximum values have also been identified for each component age group using the historical mortality data at the Indian sub-national level, which shall serve as benchmark values to be used and referred to in future academic and research endeavors.

## Conclusions

An overall reduction in the mortality rates of rural Indian women has been observed over the years in India. The success of public health interventions to reduce under five mortality rate is evident as the female rural mortality rates have reduced sizably for all states. However, there is still significant scope for improvement for all states in terms of reduction in mortality rates for other component age groups. Further, there is a need to divert attention toward the female geriatric (60+ years) population as the mortality rates are still high. Based on the incremental performance, the gap between the top and bottom performing states in term of feinequalities mortality rates in the rural areas is mainly attributed to multiple factors emerging from literature like, immunization coverage, the proportion of registered Antenatal checkups (ANCs), institutional deliveries, total fertility rate, the number of healthcare providers, presence of dedicated health workforce and alike. Robust policies and interventions should be developed at the state level to identify and address the state-specific determinants, which will help in lowering the rural female mortality rates at the national level.

## Data Availability

Data is held in a public repository:- (https://data.mendeley.com/datasets/v5gskzj78x/1) DOI- 10.17632/v5gskzj78x.1

## Acknowledgements

Not applicable

## Supporting information-

**S1 Table: Minimum and maximum benchmark values of Age-Specific Mortality Rates**

**S2 Map: Mapping of top five and bottom five states based on composite Age-Specific Mortality index scores in base year (2011)**

**S3 Map: Mapping of top five and bottom five states based on composite Age-Specific Mortality index scores in reference year (2018)**

## Notes

### Competing Interest Statement

The authors have declared no competing interest.

### Funding Statement

The author(s) received no specific funding for this work.

### Author Declarations

The study is purely based on secondary data, which is freely available to the general public. The study does not involve any interaction with the participants/human subjects. Hence, ethical approval was not required for this study.

